# Telehealth consultation in medical education: a mixed methods study of medical students’ and educators’ experiences

**DOI:** 10.64898/2025.12.10.25342002

**Authors:** Lisa-Christin Wetzlmair-Kephart, Andrew O’Malley, Veronica O’Carroll

## Abstract

**Background:** The COVID-19 pandemic accelerated the adoption of telehealth in healthcare delivery and education. However, the integration of telehealth into medical curricula is often limited to institutional initiatives and remains scattered. This raises questions about the experienced barriers and enablers for education in telehealth consultation.

**Objective:** This study sought to explore medical students’ and educators’ experiences with telehealth consultations in academic and clinical learning environments during the pandemic in the United Kingdom.

**Methods:** A cross-sectional mixed-methods design was employed. Quantitative data was gathered via online questionnaires, while qualitative insights were derived from semi-structured interviews. A total of 248 participants (153 students, 87 educators) responded to the survey, of those, 23 participants (13 students, 10 educators) were interviewed.

**Results:** Telehealth consultations were primarily taught and delivered during clinical placements. The pandemic and subsequent educational adjustments were identified as enabling factors. Exposure to telehealth increased students’ confidence and readiness, particularly through active participation in consultations. Barriers included inadequate technical infrastructure, lack of confidential spaces, and limited confidence of educators teaching telehealth consultation.

**Conclusion:** The findings highlight the need for an integrated telehealth curriculum that incorporates practical exposure and addresses technical limitations in both universities as well as healthcare settings.

## Introduction

The developments and increased use of information and communication technology in person-centred health and social care has allowed new and potentially faster, more interprofessional workflows [1]. Telehealth enables healthcare professionals to deliver, and patients and care givers to receive, care remotely, closer to the patients’ living environment. Under the premise of available and adequately used telehealth infrastructure [2], the use of technology in healthcare has highlighted benefits including: increased access to timely care [3], patient satisfaction [4,5], and cost savings [6]. Conversely, numerous downsides of technology-assisted care have been reported such as impeded access to telehealth services for some population groups [7–9]. Patients express worries about being less seen as human and more like a name to be ticked off a calling list [10,11]. Healthcare professionals perceive it harder to build rapport with their patients, and worry about introducing barriers to accessible care for disadvantaged population groups such as persons with limited digital health literacy or access to technology and internet [12,13].

Healthcare professionals acknowledge the differences between in-person and remote care and the required adaptations in clinical reasoning and technical and non-technical skills (e.g. physical examination, communication) [14]. Nevertheless, the exposure of healthcare professionals – especially medical doctors – to telehealth training or education is limited, with a significant percentage of professionals reporting ad-hoc training with the onset of the COVID-19 pandemic [15,16]. The outbreak of COVID-19 accelerated the awareness of opportunities that remote care can offer. Similarly, medical education followed the imposed social distancing and lockdown, leading to students participating in telehealth simulations and clinical experiences out of necessity [17]. Even though medical doctors have identified training as essential to ensure patient-safety and valuable for facilitating a transition from in-person to remote care [16], telehealth education still seems to neither be fully embedded in pre- or post-qualification education [18–20], and often depends on universities’ and faculty’s initiatives [21]. Remote consultations rates peaked during the pandemic but slightly decreased since 2022 [22,23]. Similarly, rapid developments of virtual reality (VR) and artificial intelligence (AI) seem to advance the conversation about telehealth consultation in clinical practice and education [24–26].

Given the developments during the COVID-19 pandemic, medical students’ and educators’ experiences of telehealth delivery should not be forgotten to ensure safe and high-quality care delivery in an increasingly digitalised healthcare system. These insights can help the medical and educational community understand learners’ as well as facilitators’ needs, and support the development of sustainable, integrated telehealth education. Therefore, the aim of this study was to illustrate medical students’ and educators’ experiences of telehealth consultations in the United Kingdom (UK) during the pandemic. Specifically, the research questions were: How was telehealth education implemented in medical education during COVID-19 and what barriers and facilitators with implementing telehealth education were experienced by medical students and educators?

This study was part of a larger PhD project which aimed to identify factors that influence the development of a telehealth curriculum in UK undergraduate medical education. As such, opinions of medical educators and students on telehealth education were investigated and previously reported [27].

## Methods

This cross-sectional mixed methods study followed a convergent design which allowed the simultaneous collection of quantitative and qualitative data to inform one another. Data analyses were performed separately and integrated again during the data interpretation phase. Quantitative data was collected using an online questionnaire comprising of a total of 29 single- and multiple-choice, and open-ended questions. The questionnaire was administered through Qualtrics [28] and developed through a test-retest approach with five medical students and five educators before being disseminated. After completing the questionnaire, participants were invited to qualitative follow-up semi-structured interviews over Microsoft Teams [29]. The topic guide for the interviews and the questionnaire can be found in the Supplementary Material.

All undergraduate medical students and educators across the United Kingdom (UK) were eligible to participate in this study. Recruitment started in May 2021; the last interview took place in March 2022. Through cluster, snowball, and convenience sampling techniques, participants were recruited via all 42 accredited medical schools in the UK. A more detailed description of the methods, including the development and administration of the research instruments, has been reported elsewhere [27].

Quantitative data was analysed descriptively reporting the total and relative number of responses. All analyses were stratified by the participants role in medical education (i.e., medical student or educator). After reviewing the raw data, two respondents who identified their role in medical education as “other” were recoded to “educator”. This was based on their provided specifications in the open text field. Further subgroup and within-group analyses (stratified based on students and educators) were omitted due to the limited response rates that would allow a meaningful interpretation of the data. The quantitative data analysis was performed in SPSS, version 26 [30], and Microsoft Excel [31].

The interviews were transcribed verbatim and together with the open-ended questions from the questionnaire analysed inductively based on the principles of thematic analysis [32,33] by the first author. To ensure the reliability of the data analysis, inter-coder-reliability was performed for a subset of 12 interview transcripts by two other researchers (AOM, VOC). Rates below 80% agreement were discussed among the researchers and necessary adaptations to the codebook were performed to increase the reliability score [34]. Data analysis was conducted in NVivo [35].

Ethics approval was obtained from the [*University omitted for peer review*] (approval code [*omitted for peer review*]). Additional approval and permission to undertake the research were sought from the Medical Schools Council (MSC). Participants’ informed consent was collected on the first page of the questionnaire. Interview participants provided their written consent via email and were asked to repeat their consent verbally before the recording started.

## Results

A total of 248 questionnaire responses were recorded between May 2021 and March 2022. 153 responses (63.8%) from medical students and 87 responses (36.3%) from educators were included in the analysis. Eight responses were excluded due to incomplete responses. Between June 2021 and March 2022, 23 follow-up interviews were conducted with 13 medical students and 10 educators. Table 1 shows the participants’ demographics. The thematic analysis identified a total of 4 themes inductively.

**Table 1:** Demographics of questionnaire respondents. Note that some totals do not equal 100% due to missing responses.

|  | Educators (n=87, 36.3%) |  |  | Students<br>(n=153, 63.8%) |
| --- | --- | --- | --- | --- |
|  | Academia<br>(n=21,<br>8.8%) | Both<br>(n=40,<br>16.7%) | Clinical<br>(n=26, 1.8%) |  |
| Gender, n (%) |  |  |  |  |
| Female | 10 (4.2%) | 21 (8.8%) | 16 (6.7%) | 118 (49.2%) |
| Male | 10 (4.2) | 17 (7.1%) | 9 (3.8%) | 31 (12.9%) |
| Affiliated University, n (%) |  |  |  |  |
| England | 10 (11.5%) | 14 (16.1%) | 7 (8.1%) | 91 (59.5%) |
| Northern Ireland | 0 | 1 (1.2%) | 1 (1.2%) | 0 |
| Scotland | 7 (8.1%) | 17 (19.5%) | 16 (18.4%) | 53 (34.6%) |
| Wales | 2 (2.3%) | 2 (2.3%) | 1 (1.2%) | 0 |
| England and Scotland | 1 (1.2%) | 0 | 1 (1.2%) | 0 |
| **Education, n (%)** |  |  |  |  |
| Bachelor's Degree | 1 (0.4%) | 5 (2.1%) | 10 (4.2%) | 48 (20.0%) |
| College / Further Education | 1 (0.4%) | 0 | 0 | 39 (16.3%) |
| High School | 0 | 0 | 0 | 48 (20.0%) |
| Master's Degree | 7 (2.9%) | 13 (5.4%) | 7 (2.9%) | 13 (5.4%) |
| Post-Graduate Diplomas | 0 | 2 (0.8%) | 1 (0.4%) | 1 (0.4%) |
| MD | 1 (0.4%) | 6 (2.5%) | 5 (2.1%) | 0 |
| PhD | 8 (3.3%) | 7 (2.9%) | 2 (0.8%) | 3 (1.3%) |
| **Year of Study, n (%)** |  |  |  |  |
| Year 1 |  |  |  | 36 (23.5%) |
| Year 2 |  |  |  | 38 (24.8%) |
| Year 3 |  |  |  | 23 (15.0%) |
| Year 4 |  |  |  | 31 (20.3%) |
| Year 5 |  |  |  | 11 (7.2%) |
| Year 6 |  |  |  | 6 (3.9%) |
| Intercalating |  |  |  | 3 (2.0%) |

### Quantitative Results

In relation to the first research question, on how telehealth education was implemented in medical education during the COVID-19 pandemic, the following results explore prior (before March 2020) and current (during the time of data collection) experiences to telehealth and learning environment and modalities.

Most students experienced telehealth consultations for the first time during COVID-19. Approximately half of the students reported that they first learned about telehealth consultations in clinical placements (n=75, 49.0%) and university settings (n=84, 54.9%) during the pandemic. As shown in Table 2, 46 students (30.1%) and 42 students (27.5%) have never experienced any form of education in clinical nor university settings, respectively.

**Table 2:** Students’ and educators’ experience with telehealth consultation. Note that some totals do not equal100% due to missing responses. Before/During refers to before and/or during the COVID-19 pandemic.

|  | Before | During | Before and during | None (%) |
| --- | --- | --- | --- | --- |
|  | n (%) | n (%) | n (%) | n |
| Have you learned about telehealth consultation techniques ... (students n=153) |  |  |  |  |
| ... in your **clinical** placements? | 5 (3.3) | 75 (49.0) | 13 (8.5) | 46 (30.1) |
| ... in a **university** setting? | 7 (4.6) | 84 (54.9) | 6 (3.9) | 42 (27.5) |
| Have you **offered** telehealth consultation in health care? (educators n=87) | 9 (10.3) | 31 (35.6) | 17 (19.5) | 20 (23.0) |
| Have you **taught** telehealth consultation techniques at university? (educators n=87) | 2 (2.3) | 22 (25.3) | 6 (6.9) | 47 (54.0) |

More than half of educators (n=47, 54.0%) did not teach telehealth consultation before nor during the COVID-19 pandemic. Most of those educators are working in the healthcare setting (n=18, 20.7%) or both an academic and the healthcare setting (n=17, 19.5%). Only two educators (2.3%) have offered telehealth consultation education prior to and six (6.9%) before and during the pandemic.

Telehealth consultations seemed to be offered at universities either throughout all years or in the second half of the studies (see Table 3). Interestingly, most students and educators did not know when telehealth consultation was taught at their universities or whether the education was mandatory for students to attend.

**Table 3:** Learning about telehealth consultations at universities: embeddedness in the medical curriculum. Note that some totals do not equal 100% due to missing responses.

|  |  | **Educators** | **Students** | **Total** |
| --- | --- | --- | --- | --- |
|  |  | **n (%)** | **n (%)** | **n (%)** |
| **When** do students learn about telehealth consultation at your university? |  | **58 (36.0)** | **103 (64.0)** | **161 (100)** |
|  | Don't know | 32 (19.9) | 38 (23.6) | 70 (43.5) |
|  | First half of the programme | 5 (3.1) | 14 (8.7) | 19 (11.8) |
|  | Second half of the programme | 11 (6.8) | 20 (12.4) | 31 (19.3) |
|  | Throughout the whole programme | 10 (6.2) | 31 (19.3) | 41 (25.5) |
| **Is telehealth consultation education at your university mandatory or voluntary?** |  | **58 (36.0)** | **103 (64.0)** | **161 (100)** |
|  | Don't know | 37 (23.0) | 36 (22.4) | 73 (45.3) |
|  | Mandatory | 12 (7.5) | 46 (28.6) | 58 (36.0) |
|  | Voluntary | 3 (1.9) | 6 (3.7) | 9 (5.6) |
|  | A combination of both | 6 (3.8) | 15 (9.3) | 21 (13.0) |

Students’ responses in relation to learning environment and modalities indicate that learning took place in simulated environments (55 responses, 4.5%) and clinical environments (58 responses, 4.7%). Educators either did not know how students learn about telehealth consultations or identified clinical placements as the main way of students learning about telehealth consultations (25 responses each, 3.6%). Both students (68 responses, 5.0%) and educators (24 responses, 3.1%) identified the main content of their education as the adequate use of telehealth consultation. The free-text answers indicate that the content was mostly referring to history-taking and communication skills. The findings are listed in Table 4.

**Table 4:** Learning about telehealth consultations at universities: Learning environment and content (multi-select responses)

|  |  | **Educators** | **Students** | **Total** |
| --- | --- | --- | --- | --- |
|  |  | **n (%)** | **n (%)** | **n (%)** |
| **How** do students learn about telehealth consultation at your university? | Clinical placement | 25 (3.6) | 58 (4.8) | 83 (4.3) |
|  | Don't know | 25 (3.6) | 18 (1.5) | 43 (2.2) |
|  | Lecture | 8 (1.2) | 31 (2.5) | 39 (2.0) |
|  | Other | 5 (0.7) | 10 (0.8) | 15 (0.8) |
|  | Self-directed | 11 (1.6) | 19 (1.6) | 30 (1.6) |
|  | Simulated environment | 21 (3.0) | 55 (4.5) | 76 (4.0) |
|  | Small groups | 14 (2.0) | 42 (3.4) | 56 (2.9) |
|  | **Total clicks** | **696** | **1224** | **1920** |
| **What** do students learn about telehealth consultation at your university? | Computer Science | 4 (0.5) | 2 (0.2) | 6 (0.3) |
|  | Don't know | 30 (3.8) | 12 (0.9) | 42 (1.9) |
|  | Ethics | 17 (2.2) | 40 (2.9) | 57 (2.6) |
|  | No teaching at all | 1 (0.1) | 19 (1.4) | 20 (0.9) |
|  | Other | 5 (0.6) | 3 (0.2) | 8 (0.4) |
|  | Research opportunities | 4 (0.5) | 7 (0.5) | 11 (0.5) |
|  | Start-ups | 3 (0.4) | 3 (0.2) | 6 (0.3) |
|  | Usage | 24 (3.1) | 68 (4.9) | 92 (4.3) |
|  | **Total clicks** | **783** | **1377** | **2160** |

Table 5 explores the second research question in relation to experienced difficulties in telehealth teaching and learning. A third of the students indicated that they experienced some technical difficulties (n=54, 35.3%) and organisational difficulties (n=53, 34.6%) when practicing telehealth consultations during university or clinical learning experiences.

**Table 5:** Technical and organisational difficulties experienced with telehealth consultation services. Note that some totals do not equal 100% due to missing responses.

|  | Technical Difficulties | Organisational Difficulties |
| --- | --- | --- |
|  | n (%) | n (%) |
| **Students (n=153)** |  |  |
| No, never | 16 (10.5) | 24 (15.7) |
| Yes, often | 19 (12.4) | 12 (7.8) |
| Yes, sometimes | 54 (35.3) | 53 (34.6) |
| **Educators (n=87)**<br>(difficulties in use experienced in a healthcare setting) |  |  |
| No, never | 3 (3.5) | 11 (12.6) |
| Yes, often | 8 (9.2) | 8 (9.2) |
| Yes, sometimes | 37 (42.5) | 29 (33.3) |
| **Educators (n=87)**<br>(difficulties in use experienced in a teaching setting) |  |  |
| No, never | 1 (1.2) | 6 (6.9) |
| Yes, often | 3 (3.5) | 5 (5.8) |
| Yes, sometimes | 24 (27.6) | 17 (19.6) |

Educators in healthcare settings experience more technical (n=37; 42.5%) and organisational difficulties (n=29; 33.3%) than educators in teaching settings (n=24, 27.6% and n=17; 19.5%, respectively).

### Qualitative Results

The sequential mixed-methods design allowed a deeper insight from the quantitative results into how telehealth consultation was implemented in medical education. The following themes were identified in the interviews and open-text responses of the questionnaire: (a) COVID-19 accelerated change and (b) Educational adjustments. Themes in relation to the second research question about experienced facilitators and barriers were (c) Exposure increases confidence and (d) Technical, structural, and organisational barriers.

#### COVID-19 accelerated change

The COVID-19 pandemic was highlighted as a significant event that prompted educators to divide clinical practice and medical education into ‘before COVID and now’ (Educator (both settings), interview 18). Before the pandemic, most students would not experience formal teaching at universities or in their placements. Medical education practice was reflecting the dominant mode of care delivery: in-person appointments and occasional telephone follow-ups.

> *… but that [telehealth consultations] wasn’t something I delivered up front. It was something that would often come out of you know the experience and the students were asking questions. – Educator (both settings), interview 2*

> *I’ve also done quite a number of telephone clinics over the last year in particular. I wasn’t doing telephone clinics before COVID. – Educator (healthcare), interview 1*

> *… before that [lockdowns] we had mostly in-person communication skills teaching. So, in years one to three you’d go into a room and there will be your peers sitting with you and then you have a mock patient and a tutor, and you do it. – Student, interview 13*

#### Educational adjustments

Once the COVID-19 lockdowns were implemented, educators were required to adjust both their clinical and educational practices. Changes in educational practices was experienced in three areas: teaching modalities, content, and clinical placements.

Adjustments to teaching modalities were necessary as in-person teaching was widely suspended during lockdowns. Technology that was available before was then not only incorporated into teaching (i.e., online learning) but also used as a medium to teach clinical skills (i.e., telehealth).

> *This is really since COVID, when we had Teams and we thought, yes, we can actually use this now for telephone consultations and video. I’m really pleased with the outcomes. It worked so well. – Educator (academia), interview 6*

The learning content for students shifted to account for both, online learning and telehealth encounters in placements. Students were introduced to aspects on how to safely perform remote physical examinations and communication.

> *… It shows the medical students how you can examine someone without physically being there … so, at the end of the day it’s a different way of teaching, I suppose. – Educator (healthcare), interview 19*

Adapting clinical placements was either necessary due to lockdown restrictions or a result of changed clinical practices. Hence, students experienced their placements either fully remote especially during the first weeks of the pandemic, or had hybrid placements, where some patients would be consulted remotely, and some were asked to come into the clinic.

> *… in first year just because we were able to see patients, and I think at that stage it just was the most natural thing to do to see them in person uhm but yeah, with COVID obviously everything changed and I had all my placements all virtual … – Student, interview 22*

#### Exposure increases confidence

Students and educators shared how exposure to telehealth can increase their confidence in conducting remote consultations. Some educators seemed to rely on their clinical and teaching experiences to cope with the changes in remote care delivery such as physical examination and communication.

> *I’ll give him [a patient] a call … and as I was doing that, I realised I would never have done that in the past I would never have thought, you know, it’s two o’clock, patient hasn’t turned up, you know what I’ll call them. … Because in a sense you didn’t do telephone consultations. That just wasn’t a thing … you didn’t initiate telephone consultations like that, whereas now you were, and it seemed a very small step to then phone somebody. – Educator (both settings), interview 2*

> *I think what I would say is that being an experienced GP makes teleconsultations very easy because I can draw on my face-to-face experience. – Educator (both settings), interview 8*

For some students who were unable to continue their in-person clinical experiences, exposure to remote consultations during their university training and clinical placements enabled them to feel more prepared to practice telehealth consultations.

> *It is helpful to see doctors take histories via phone and determining when you have to get the patient to come in instead of just having a phone consultation. I think it’s always quite difficult, but if you see lots of lots of doctors calling then you can kind of understand when it’s important to get them to come in. I’ve seen quite a bit of that which has been helpful. – Student, interview 9*

> *There was a lot of phone consultations … and I was doing some of them by myself, but I got a chance to learn through observing more than learn through official teaching by the GPs. I would watch them do it and then they give me a go with the second one or something and then we get like feedback on how I did. – Student, interview 13*

#### Technical, structural, and organisational barriers

Quality and functionality of technology available in the UK National Health Service (NHS) seemed to hinder positive educational experiences for both students and educators.

> *technical difficulties interrupting teleconsultations – Student, questionnaire response 40*

> *Devices and general computer quality in the NHS is poor, making telecommunications more difficult – Student, questionnaire response 128*

> *Because it would just be a telephone and their speaker system wasn’t working so I all I could hear was the GP speaking and I couldn’t hear the other side of the conversation, so that was where it was a bit of a disadvantage having the teleconsultations style, it’s not so easy if you’re doing it in a teaching setting, if you just using a telephone – Student, interview 22*

Interestingly, only students conveyed the impression that telehealth consultation might require the acquisition of different skills to the expense of other competencies. In combination with experienced technical difficulties, one student shared their frustration with learning.

> *Some of the clinics would have a couple of phone calls in them and you’d get to see that happen occasionally and annoyingly without actually being able to hear the patient. … That teaches you nothing. – Student, interview 14*

Educators perceived structural barriers such as limited physical, confidential spaces in the NHS as a limitation for both conducting and teaching telehealth consultations. It appears that with the rapidly required increased use of technology, professional exchange of good practices in telehealth consultations are more challenging.

> *The clinical sites are set up incorrectly to provide a lot of telemedicine. Large clinic rooms are needed to examine patients etc. But if everyone is doing telemedicine, we could use smaller rooms with PCs in them. I often find it difficult to find a place where I can work because there is no physical room to access PCs to phone all the patients. – Educator (healthcare), questionnaire response 208*

> *I suppose I feel I’m working in a silo because I’m the only one who’s doing it [telehealth consultation] in my unit. Compared to other departments they’re not really using it as much. – Educator (healthcare), interview 19*

> *The challenges I guess for me, we don’t really know what other departments and things do, so it would be useful to be able to share knowledge with other people and see how they are consulting and how they’re doing it because we’ve all kind of individually adapted to it. – Educator (both settings), interview 17*

## Discussion

The findings of this study identified enablers and barriers in telehealth consultation training in UK undergraduate medical education. Rapid changes during the COVID-19 pandemic allowed broad exposure to telehealth consultation for both students and educators. A large percentage of students could not recall any education on telehealth consultations, and a larger percentage of educators claimed never to have taught telehealth consultations. If students were exposed to these remote consultations, they often learned about it in their clinical placements or simulated learning environments; however, many participants did not know of any telehealth education taking place in their affiliated universities. Inadequate technical equipment limited the deployment of telehealth education, leading to a diminished interest in pursuing this mode of clinical practice.

Medical education rapidly adopted telehealth consultations in both academic and clinical learning environments with the onset of the pandemic. One of the issues that emerged from this transition from in-person to remote care, was the unprecedented, fast, and unforeseen change and the limited structural and organisational preparedness. Educators as well as students perceived a level of disengagement in students’ learning. Students described themselves as passive because it was hard for the educators to involve them into remote consultation processes [36]. Educators were faced with limited confidential, quiet places for telehealth consultations, which negatively impacted students’ education [36], as well as providing telehealth to patients [37]. Interestingly, placements during the pandemic, where students were consulting with patients from their own home or university environments (as opposed to clinical environments), did not seem to encounter those structural barriers [38–40]. However, in those remote placements, students highlighted that communication with their supervisors was sometimes slow and cumbersome [38]. Nevertheless, international experiences from Australia, Brazil, and the USA widely indicate that telehealth education is well-received by medical students [36,38–40], given students can actively engage and initiate the patient encounters. De-briefing sessions with educators after telehealth consultations seemed to enhance students’ perception on impactful telehealth learning opportunities [38,40]. Hence, the integration of students into remote clinical workflows seems to be feasible, even during rapid required adaptations that can cause stress and confusion as reported in this study. This might imply that the future of telehealth consultation in medical education requires careful consideration of structural, administrative, as well as technical solutions in clinical settings to develop a fully engaging and impactful telehealth education [37].

Studies undertaken in academic, pre-clinical learning environments further illustrate the importance and feasibility of telehealth consultation education. In line with the findings of this study, competencies of educators in telehealth are perceived as essential to adequately train students and ultimately improve the learning experience for students [36]. However, the sole clinical exposure might not be sufficient to enhance students’ confidence in their telehealth skills [38]. Telehealth competency frameworks have been developed to support telehealth education for healthcare professionals [24–26]. Even comprehensive frameworks that include digital health aspects such as health information systems and health data science, seem to have one thing in common: ultimately increase students’ competencies and overall understanding of the safe, person-centred, ethical implementation of telehealth consultation into clinical workflows [26,41,42]. Acknowledging the limitations of telehealth consultations and skills acquisition such as procedural skills and physical examinations [43], the value of educating students in the fundamentals of telehealth in academic learning environments cannot be neglected. Telehealth taught by educators with experience in providing remote care has the potential to ensure that students can assess the impact of technologies on their patients’ healthcare needs [36,44]. Furthermore, knowledge about legal regulations and ethical considerations can prevent medical malpractice [44] and subsequently ensure patients’ privacy and the protection of their data.

With the awareness of limited telehealth consultation training for students as well as educators and practitioners prior to the pandemic [15], this study is another piece to further advocate for integrated telehealth education during undergraduate, postgraduate education as well as continuous professional development training.

It is also worth noting that the focus of this study was solely on medical students rather than healthcare professionals in general. Evidence from nursing and allied health professionals [45,46] demonstrates promising initiatives to integrate telehealth into their curricula, suggesting that broader inclusion and comparison may provide a more comprehensive understanding of telehealth education across different healthcare disciplines.

## Conclusion

Telehealth consultation represents a pivotal opportunity in modern care delivery, thus requiring integration into medical education. To optimise telehealth consultation in academic and clinical learning and subsequently in clinical practice, a structured integration of telehealth competencies, technical preparedness, and practical exposure is vital. Empowering educators with the tools to lead impactful training and embedding telehealth into curricula can ensure future healthcare professionals are equipped to deliver ethical, effective, and person-centred remote consultations and care.

## Data Availability

Data are available from the University of St Andrews Research Repositry under: https://doi.org/10.17630/84eb74f3-e316-4618-a112-19b2f24377ac The data set will be made availableon 9 May 2028.

## References

1. Downes E, Horigan A, Teixeira P. The Transformation of Health Care For Patients: Information and Communication Technology, Digiceuticals, and Digitally Enabled Care. Journal of the American Association of Nurse Practitioners. 2019;31: 156–161. doi:10.1097/JXX.0000000000000109

2. Kobeissi MM, Hickey JV. An Infrastructure to Provide Safer, Higher-Quality, and More Equitable Telehealth. The Joint Commission Journal on Quality and Patient Safety. 2023;49: 213–222. doi:10.1016/j.jcjq.2023.01.006

3. Charalambous J, Hollingdrake O, Currie J. Nurse Practitioner Led Telehealth Services: A Scoping Review. Journal of Clinical Nursing. 2024;33: 839–858. doi:10.1111/jocn.16898

4. Hamiel U, Eshel Fuhrer A, Landau N, Reches A, Ponger P, Elhanan E, et al. Telemedicine Versus Traditional In-Person Consultations: Comparison of Patient Satisfaction Rates. Telemedicine and e-Health. 2024;30: 1013–1019. doi:10.1089/tmj.2023.0273

5. Kruse CS, Krowski N, Rodriguez B, Tran L, Vela J, Brooks M. Telehealth and Patient Satisfaction: A Systematic Review and Narrative Analysis. BMJ Open. 2017;7: e016242. doi:10.1136/bmjopen-2017-016242

6. De Guzman KR, Gavanescu D, Smith AC, Snoswell CL. Economic Evaluations of Telepharmacy Services in Non-Cancer Settings: A Systematic Review. Research in Social and Administrative Pharmacy. 2024;20: 246–254. doi:10.1016/j.sapharm.2024.01.002

7. Kalicki AV, Moody KA, Franzosa E, Gliatto PM, Ornstein KA. Barriers to Telehealth Access Among Homebound Older Adults. Journal of the American Geriatrics Society. 2021;69: 2404–2411. doi:10.1111/jgs.17163

8. Ng BP, Park C, Silverman CL, Eckhoff DO, Guest JC, Díaz DA. Accessibility and Utilisation of Telehealth Services Among Older Adults During COVID-19 Pandemic in the United States. Health & Social Care in the Community. 2022;30. doi:10.1111/hsc.13709

9. Shang DR, Williams C, Baynard C, Joshi A, Pitisci J, Saldanha S. Exploring the Interplay Between Digital Divide, Health Resource Shortage, and Telehealth Utilization. Association for Information Systems Electronic Library. Paphos, Cyprus: AIS TREO Papers; 2024. pp. 1–5. Available: https://aisel.aisnet.org/treos_ecis2024/33

10. Gordon HS, Solanki P, Bokhour BG, Gopal RK. “I’m Not Feeling Like I’m Part of the Conversation” Patients’ Perspectives on Communicating in Clinical Video Telehealth Visits. Journal of General Internal Medicine. 2020;35: 1751–1758. doi:10.1007/s11606-020-05673-w

11. Grīnfelde M. Face-to-Face with the Doctor Online: Phenomenological Analysis of Patient Experience of Teleconsultation. Human Studies. 2022;45: 673–696. doi:10.1007/s10746-022-09652-4

12. Alashek W, Ali S. Satisfaction With Telemedicine Use During COVID-19 Pandemic in the UK: A Systematic Review. Libyan Journal of Medicine. 2024;19: 2301829. doi:10.1080/19932820.2024.2301829

13. Jonasdottir SK, Thordardottir I, Jonsdottir T. Health Professionals’ Perspective Towards Challenges and Opportunities of Telehealth Service Provision: A Scoping Review. International Journal of Medical Informatics. 2022;167: 104862. doi:10.1016/j.ijmedinf.2022.104862

14. Curran V, Hollett A, Peddle E. Training for virtual care: What do the experts think? DIGITAL HEALTH. 2023;9: 1–8. doi:10.1177/20552076231179028

15. Garber K, Gustin T. Telehealth Education: Impact on Provider Experience and Adoption. Nurse Educator. 2022;47: 75–80. doi:10.1097/NNE.0000000000001103

16. Walley D, McCombe G, Broughan J, O’Shea C, Crowley D, Quinlan D, et al. Use of Telemedicine in General Practice in Europe Since the COVID-19 Pandemic: A Scoping Review of Patient and Practitioner Perspectives. Lichtner V, editor. PLOS Digital Health. 2024;3: e0000427. doi:10.1371/journal.pdig.0000427

17. Muntz MD, Franco J, Ferguson CC, Ark TK, Kalet A. Telehealth and Medical Student Education in the Time of COVID-19 - and Beyond. Academic Medicine. 2021;96: 1655–1659. doi:10.1097/ACM.0000000000004014

18. Cassiday OA, Nickasch BL, Mott JD. Exploring Telehealth in the Graduate Curriculum. Nursing Forum. 2021;56: 228–232. doi:10.1111/nuf.12524

19. Chike-Harris KE, Durham C, Logan A, Smith G, DuBose-Morris R. Integration of Telehealth Education into the Health Care Provider Curriculum: A Review. Telemedicine and e-Health. 2021;27: 137–149. doi:10.1089/tmj.2019.0261

20. Hilty DM, Torous J, Parish MB, Chan SR, Xiong G, Scher L, et al. A Literature Review Comparing Clinicians’ Approaches and Skills to In-Person, Synchronous, and Asynchronous Care: Moving Toward Competencies to Ensure Quality Care. Telemedicine and e-Health. 2021;27: 356–373. doi:10.1089/tmj.2020.0054

21. Sharkiya SH. Telehealth Challenges in Nursing: A Narrative Review. International Journal of Clinical Science and Medical Research. 2024;04: 287–290. doi:10.55677/IJCSMR/V4I8-01/2024

22. Howlett O, Harnetty T, Barrett S. Maintaining the Use of Telehealth For Delivering Rehabilitation Services in a Regional Hospital post-COVID19: Learning From Telehealth Delivery Rates and Staff Experiences. Clinical Rehabilitation. 2025;39: 679–689. doi:10.1177/02692155251326050

23. Vestesson EM, De Corte KLA, Crellin E, Ledger J, Bakhai M, Clarke GM. Consultation Rate and Mode by Deprivation in English General Practice From 2018 to 2022: Population-Based Study. JMIR Public Health and Surveillance. 2023;9: e44944. doi:10.2196/44944

24. Al Baalharith IM, Aboshaiqah AE. A Delphi Study on Identifying Competencies in Virtual Healthcare for Healthcare Professionals. Healthcare. 2024;12: 739–749. doi:10.3390/healthcare12070739

25. Car J, Ong QC, Fox TE, Leightley D, Kemp SJ, Švab I, et al. The Digital Health Competencies in Medical Education Framework. JAMA Network Open. 2025;8: e2453131. doi:10.1001/jamanetworkopen.2024.53131

26. Jacob MFA, Fandim JV, Reis FJJ, Hartvigsen J, Ferreira PH, Saragiotto BT. Defining Core Competencies For Telehealth in Healthcare Higher Education: A Delphi Study. Musculoskeletal Science and Practice. 2025;75: e103244. doi:10.1016/j.msksp.2024.103244

27. Wetzlmair-Kephart L-C, O’Malley A, O’Carroll V. Medical Students’ and Educators’ Opinions of Teleconsultation in Practice and Undergraduate Education: A UK-Based Mixed-Methods Study. PLoS ONE. 2025;20: e0302088. doi:10.1371/journal.pone.0302088

28. Qualtrics. Qualtrics. 2025 [cited 1 Nov 2025]. Available: https://www.qualtrics.com/uk/

29. Microsoft. Microsoft Teams. 2025. Available: https://www.microsoft.com/en/microsoft-teams/group-chat-software

30. Corp IBM. IBM SPSS Statistics for Windows. IBM; 2019. Available: https://www.ibm.com/uk-en

31. Microsoft. Microsoft Excel. 2025. Available: https://office.microsoft.com/excel

32. Braun V, Clarke V. Successful qualitative research a practical guide for beginners. London: SAGE Publications; 2013.

33. Guest G, MacQueen KM, Namey EE. Applied Thematic Analysis. London: SAGE Publications; 2012. doi:10.4135/9781483384436

34. Fereday J, Muir-Cochrane E. Demonstrating Rigor Using Thematic Analysis: A hybrid Approach of Inductive and Deductive Coding and Theme Development. International Journal of Qualitative Methods. 2006;5: 80–92. doi:10.1177/160940690600500107

35. Lumivero. NVivo. 2023. Available: https://lumivero.com/products/nvivo/

36. Pit SW, Velovski S, Cockrell K, Bailey J. A Qualitative Exploration of Medical Students’ Placement Experiences With Telehealth During COVID-19 and Recommendations to Prepare Our Future Medical Workforce. BMC Medical Education. 2021;21: 431–444. doi:10.1186/s12909-021-02719-3

37. Meng G, McAiney C, Perlman CM, McKillop I, Tisseverasinghe T, Chen HH. Service Process Factors Affecting Patients’ and Clinicians’ Experiences on Rapid Teleconsultation Implementation in Out-Patient Neurology Services During COVID-19 Pandemic: A Scoping Review. BMC Health Services Research. 2022;22: 534–551. doi:10.1186/s12913-022-07908-4

38. Kopp AR, Rikin S, Cassese T, Berger MA, Raff AC, Gendlina I. Medical student remote eConsult participation during the COVID-19 pandemic. BMC Medical Education. 2021;21: 120–130. doi:10.1186/s12909-021-02562-6

39. Pedroso TM, Vasconcelos IM, De Amorim CL, Coelho LR, Corrêa MAM, De Aguiar VB, et al. Medical Students Experience in Working in a Public COVID-19 Telehealth Program: A Descriptive Study. BMC Medical Education. 2024;24: 756. doi:10.1186/s12909-024-05722-6

40. Weber AM, Dua A, Chang K, Jupalli H, Rizwan F, Chouthai A, et al. An Outpatient Telehealth Elective for Displaced Clinical Learners During the COVID-19 Pandemic. BMC Medical Education. 2021;21: 174–182. doi:10.1186/s12909-021-02604-z

41. Bajra R, Frazier W, Graves L, Jacobson K, Rodriguez A, Theobald M, et al. Feasibility and Acceptability of a US National Telemedicine Curriculum for Medical Students and Residents: Multi-Institutional Cross-Sectional Study. JMIR Medical Education. 2023;9: e43190. doi:10.2196/43190

42. Mariño RJ, Capurro D, Merolli M. Pilot Implementation of a Telehealth Course for Health Professions Students. BMC Medical Education. 2024;24: 963–971. doi:10.1186/s12909-024-05931-z

43. Saad S, Richmond C, King D, Jones C, Malau-Aduli B. The Impact of Pandemic Disruptions on Clinical Skills Learning for Pre-Clinical Medical Students: Implications for Future Educational Designs. BMC Medical Education. 2023;23: 364–377. doi:10.1186/s12909-023-04351-9

44. Aungst TD, Patel R. Integrating digital health into the curriculum - considerations on the current landscape and future developments. Journal of Medical Education Curriculum Development. 2020. doi:10.1177/2382120519901275

45. Anil K, Bird A, Bridgman K, Erickson S, Freeman J, McKinstry C, et al. Telehealth Training and Education for Allied Health Professionals: A Scoping Review. Telemedicine Reports. 2025;6: 76–90. doi:10.1089/tmr.2024.0083

46. Rettinger L, Putz P, Aichinger L, Javorszky SM, Widhalm K, Ertelt-Bach V, et al. Telehealth Education in Allied Health Care and Nursing: Web-Based Cross-Sectional Survey of Students’ Perceived Knowledge, Skills, Attitudes, and Experience. JMIR Med Educ. 2024;10: e51112. doi:10.2196/51112

